# Genomic Insights into Opioid Addiction: Identification of genomic variants from Gene Expression data

**DOI:** 10.1101/2025.02.22.25322710

**Authors:** Swati Ajmeriya, Biswadip Chatterjee, Subhradip Karmakar

## Abstract

**Background:** Next-generation sequencing (NGS) has transformed high-throughput DNA and RNA analysis, facilitating the rapid identification of clinically relevant genetic variants. Opioid Use Disorder (OUD), a chronic condition characterized by relapse and remission cycles, poses significant challenges for genomic investigations. While whole-exome sequencing (WES) and whole-genome sequencing (WGS) serve as robust approaches for variant detection, their high cost restricts widespread use. RNA sequencing (RNA-Seq) presents a viable alternative; however, the complexity of the transcriptome complicates reliable variant identification. Despite these challenges, RNA-Seq has emerged as a valuable tool for detecting single nucleotide polymorphisms (SNPs) in conditions with limited WES data, such as OUD.

**Methods:** In this study, RNA-Seq data from postmortem ventral midbrain specimens of chronic opioid users (PRJNA492904) were analyzed to identify variants associated with OUD. Given its established involvement in opioid addiction and impulsivity, we hypothesized that the *NRXN3* gene would harbor a significant number of variants. Variant analysis was conducted across eight genes: *BDNF, DRD2, DRD3, NRXN3, OPRD1, OPRM1,* and *NGFB*—with a primary focus on *NRXN*.

**Results:** Our results revealed that *NRXN3* exhibited the highest variant burden among the analyzed genes, highlighting its potential role in OUD pathogenesis and reinforcing its association with opioid addiction.

**Conclusion:** Our study highlights the significance of transcriptomic variant analysis in opioid addiction and underscores the potential of NRXN3 variants as biomarkers or therapeutic targets, warranting further investigation.

## Introduction

Substance use disorders are characterized by frequent relapses, with relapse rates ranging from 56.8% to 81.8%[1,2]. The World Drug Report published by the CDC shows that the number of drug overdose deaths increased by nearly 30% from 2019 to 2020 and nearly 75% of the 91,799 drug overdose deaths in 2020 involved an opioid; from 2019 to 2020, Opioid-involved death rates increased by 38%[3]. The past several decades have seen parallel increases in the prescription of opioid drugs and prescription-related opioid overdose deaths, followed by a resurgence in heroin abuse and deaths, and, most recently, there has been an abrupt rise in opioid deaths involving fentanyl or fentanyl analogs in the United States[4]. Thus, despite best and continuous efforts, the outcome of opioid use remains poor and it remains an important cause of premature death among the users. It has been established that genetic factors contribute to the vulnerability to developing drug addiction and to the effectiveness of its treatment [5,6]. Genetic factors majorly contribute to the individual variations of the risk of developing opioid addiction with ∼ 60% of the population[7,8]. Identification of these factors may increase our understanding of the disorders, help in the development of new treatments, and advance personalized medicine. Considerable effects on protein function and gene expression can be caused by variants occurring in coding regions and regulatory sequences, respectively[9]. Opioid exposure and OUD are linked to widespread gene expression changes in the brain and opioid-related variants have been reported to be associated with specific drug addictions[10,11]. The significance of utilizing human postmortem brains for molecular characterization of neuropsychiatric disorders has been thoroughly reviewed [12] This approach holds the potential to greatly enhance our understanding of psychiatric disorders.

Drugs of abuse share the property of acutely increasing the signaling of dopamine (DA)-synthesizing midbrain neurons to several forebrain targets[13] [14]. With continued drug use, neuroadaptations such as drug dependence, craving, and relapse are thought to arise from persistent changes in gene expression[13,14]. Gene expression changes have been identified in human forebrain targets of dopamine (DA) signaling, such as the nucleus accumbens and prefrontal cortex[15–17]. However, there is limited information regarding both gene expression changes and SNPs in midbrain human DA neurons, despite their crucial role in the circuitry of addiction. Today, in the era of biomedical research, the accessibility of Next-generation sequencing (NGS) offers a streamlined means to comprehensively uncover all variants including SNPs and insertion and deletion variants (indels). Several methods have been implemented for initial SNPs discovery in a high-throughput manner, such as whole-genome sequencing, exome capture, RNA sequencing, methylated DNA sequencing, and restriction enzyme (RE) digestion[18]. Recently, transcriptome sequencing, or RNA-seq, has become one of the most representative high-throughput sequence-based techniques because of its high accuracy and cost-effectiveness[9]. However, the identification of variants from existing RNA-Seq remains a challenge because of the transcriptome complexity. At present, Illumina with good sequencing coverage and read quality, is applied to many plant species for variant detection by RNA-seq[9]. Some existing studies prove the reliable and accurate identification of variants from RNA-Seq data. One such study has evaluated the accuracy of SNPs identified from RNA-Seq data that has utilized its application to produce massive SNPs at relatively low cost[19]. Furthermore, RNA-Seq has been shown to detect genomic variations in expressed regions like DNA-seq, as elucidated by[20]. Furthermore, a Trinity-GATK pipeline has been developed in a research study to identify reliable variants from RNA-Seq data [9]. These findings demonstrate the prospective benefits of utilizing RNA-Seq or transcriptome sequencing as high-throughput sequence-based technology for variant discovery. Previously, the applications and advantages of RNA-Seq to discover variants have been reviewed and described. RNA-Seq has been described to be able to discover thousands of variants as well as the expression levels of functional genes with sequence variations, all at a reasonable cost[9]. For these reasons, RNA-seq is developing into an extensive application in genetic polymorphism analysis[9]. There is a significant paucity of genomic data related to OUD and other psychiatric disorders, resulting in a limited understanding of the molecular and genetic mechanisms underlying these conditions. This research gap persists despite the established association between gene expression changes and brain physiology, particularly in opioid addiction. In the current study, we employed PRJNA492904 gene expression data to detect variants derived from ventral midbrain specimens from chronic opioid users due to the limited information on OUD-related variants. We have elucidated the significance of variants in the genes previously found to have a crucial role in OUD. The current investigation represents, to our knowledge, the first unbiased examination of variants in the midbrain associated with human opioid abuse in the expression region of the human genome. Our analyses revealed variants across seven candidate genes (BDNF, DRD2, DRD3, NRXN3, OPRD1, OPRM1, and NGFB), among which NRXN3 exhibited the highest frequency of genetic variation. Some pieces of evidence demonstrate the indispensable role of α-NRXNs in the regulation of normal neurotransmitter release mechanisms[21]. NRXNs have been considered a potential candidate gene for drug addiction[22]. In European and African American samples, 38 loci containing SNPs were strongly associated with the amount of opioid abuse, including NRXN3 [23]. Furthermore, the involvement of NRXN3 in normal neurotransmitter release and synaptic integrity suggests that variation in this gene may have widespread and substantial effects on key behavioral phenotypes such as impulsivity[24]. Therefore, by utilizing the RNA-Seq data this study examined the genetic polymorphisms in *NRXN3* relative to the other candidate genes in transcriptome expression data. The pronounced variants observed in NRXN3 support the notion that NRXN3 may be involved in mediating impulsive and addictive behavior among individuals with opioid addiction as well as the reliability of RNA-Seq to detect genetic variants.

## Material and Methods

### Acquisition of RNA-Seq data for variant calling

We used publicly available RNA-seq data on humans with opioid abuse from the Sequence Read Archive (SRA) on postmortem ventral midbrain specimens from chronic opioid users (N = 30) and drug-free control subjects (N = 20), as described in the Methods with project ID PRJNA492904[25].

### Case Inclusion and Exclusion Criteria for the dataset PRJNA492904

The inclusion criteria for the opioid abuse cohort (n=30) included: (1) a verifiable history of opioid abuse, corroborated by a toxicology report indicating the presence of opioids and a forensic analysis attributing death directly to opioid consumption. Inclusion was also extended to a subset of cases with concurrent detection of opioids and cocaine, reflecting the prevalent pattern of polydrug use with opioids. (2) Subjects with a positive toxicological screening for non-opioid substances of abuse (e.g., alcohol, cannabinoids, anxiolytics, barbiturates) were systematically excluded from participation. For the control group (n=20), the inclusion criteria were: (a) absence of a documented drug abuse history and negative toxicological tests for opiates, cocaine, and other substances of abuse or central nervous system (CNS) medications. The causes of death in control cases were primarily cardiovascular incidents or gunshot injuries. Exclusion criteria for either group encompassed: (a) a documented history of neurologic or psychiatric disorders, death by suicide, evidence of neuropathological conditions, severe chronic diseases, an estimated postmortem interval exceeding 20 hours, or biochemical indicators of compromised tissue sample integrity or extended perimortem agonal phase, such as a brain pH below 6.2 or an RNA integrity number (RIN) less than 6.0.

### Pre-processing of data

We performed the quality control analysis of the RNA-Seq raw sequence data using fastp for trimming of low-quality data, and trimming of adapters[26]. FastQC (https://www.bioinformatics.babraham.ac.uk/projects/fastqc/ ) and multiqc[27] were used to assess the quality of data.

### Alignment of reads and RNA-Seq variant identification

The mapping of RNA-Seq reads to a UCSC reference human genome hg38 reference genome was done using a STAR aligner with 2-pass mode because it increases sensitivity[28]. STAR is also recommended in GATK best practices workflow (<u>RNAseq short variant discovery (SNPs + Indels) – GATK</u> (broadinstitute.org) for the identification of short variants from RNA-Seq data and to get better alignments around novel splice junctions. From the STAR alignments’s Ist pass mode, *SJ.out.tab* was generated consisting of all the junctions obtained from the 1-pass mapping step and was used as the input for the 2^nd^ pass mode. Sorted BAM files based on genomic coordinates were obtained from the STAR output. The base quality score recalibration was done to recalibrate base quality scores. GATK HaplotypeCaller was used to find variants in the genome, including SNV and indels[29]. Picard suite was used to mark duplicates. The process of splitting reads containing N cigar string into multiple supplementary alignments, incorporating hard clips to address mismatching overhangs, was executed using the "SplitNCigarReads" function within GATK4[30]. The software tool, ANNOVAR was used to study the effect of variants on gene and protein function, for the interpretation and prioritization of single nucleotide variants and Indels[31]. SnpEff, an independent open-source platform used for annotating these variants using databases, filters, and manipulating genomic annotated variants based on their genomic locations, such as intronic, untranslated region (5′ UTR or 3′UTR), upstream, downstream, splice site, or intergenic regions[32]. SnpSift CaseControl tool was used for a case-control study of opioid(n=30) and non-opioid(n=20) subjects and filter data to find out the most significant variants[32]. dbSNP was utilized for the retrieval of genetic variants, along with the acquisition of allele population frequencies sourced from databases such as 1000 Genomes, gnomAD, dbGaP, and TOPMED, among others[33].

### Variant Prioritization

For variant prioritization, variant scores were normalized by applying the quality filtration using the "VariantFiltration" function of the Genome Analysis Toolkit (GATK), including Variant confidence normalized by unfiltered depth of variant samples QualByDepth (QD) <2, Strand bias estimated using Fisher’s Exact Test FisherStrand (FS) <60, Strand bias estimated by the symmetric odds ratio test SOR<9.0 and root mean square mapping quality over all the reads at the site MQ>40. Subsequently, variants within specified genes (BDNF, DRD2, DRD3, NRXN3, OPRD1, OPRM1, and NGFB) were extracted using the SnpSift filter function. Amino acid substitution effects on protein function were predicted using various scores including SIFT, PolyPhen, LRT, MutationTaster, MutationAssessor_pred, FATHMM_pred, fathmm_MKL_coding_pred, PROVEAN_pred, MetaSVM_pred, and MetaLR_pred sourced from the dbnsfp database[34]. Potentially deleterious non-synonymous(nsSNPs) were filtered out if they were predicted to have harmful effects by mutation annotators such as Mutation Taster, FATHMM, and PROVEAN. [26]. Further, variants were prioritized if they were exclusively present in Cases (both homozygous and heterozygous) and absent in controls. Variants affecting both coding and non-coding regions, as well as both known and novel variants, were prioritized for further analysis. Variants were classified as novel if they were absent in dbSNP database. Variants were categorized as SNP and were later sorted into missense, nonsense, and silent mutations if they were present. Utilizing the bcftools plot-vcfstats, a bar plot elucidating the number of substitutions for each gene was created (https://samtools.github.io/bcftools/bcftools.html ). Bcftools query function was utilized for counting the total number of SNPs and Indels(https://samtools.github.io/bcftools/bcftools.html ).

## Results

### RNA-Seq Variant frequency analysis

In the study aimed at identifying genetic variants associated with opioid addiction, a Case-Control analysis was conducted, comparing opioid-addicted subjects (n=30) to non-opioid subjects (n=20). Analysis using the CaseControl tool of SnpSift revealed a total of 60,13,139 annotated variants, including 3778 SNPs, 485 indels, and variants of unknown class and significance. Subsequently, variants segregating exclusively in case samples, rather than control samples, were prioritized to mitigate false positives. Further, investigation focused on variants within the NRXN3 gene and seven other genes (BDNF, DRD2, DRD3, OPRD1, OPRM1, and OPRK1 previously linked to opioid addiction[35]. We found the absence of any variant in the NGFB gene. After variant annotation with ANNOVAR and SnpFff, the variants were filtered based on predetermined thresholds, resulting in variant distributions across various genomic regions. Among the eight analyzed genes, NRXN3 exhibited the highest number of variants. Following variant filtration, the number of variants in NRXN3 decreased from 8,944 to 8,939, whereas the variant numbers for the other genes remained unchanged, as detailed in **Table I**. The variant profiles for the genes, present exclusively in the cases cohort and absent in the control group, were analyzed. The total number of variants in cases was determined both before and after applying thresholds, to evaluate the significance of these thresholds on the variants derived from the RNA-Seq data. **Fig. I** illustrates the variants identified in all seven genes, both before and following the application of thresholds as described in the Materials and Methods section. Further, to provide a comprehensive understanding, the frequency of variants present in control samples was assessed and compared with variants observed in case samples. **Fig. II** depicts the total variants identified in case and control samples. The bar graph indicates that the maximum number of variants were found in the case samples, particularly in the NRXN3 gene. This comparison aimed to elucidate the differences in variant profiles between opioid-addicted and non-opioid subjects, shedding light on potential genetic contributors to opioid addiction susceptibility. This also demonstrates the ability of the transcriptome data to identify the variations accurately. We further present a detailed analysis of the variant profiles for each gene. In this study we designate variants rsId with known varaints and the varaints with absence of rsId with novel variants.

**Table. I.**
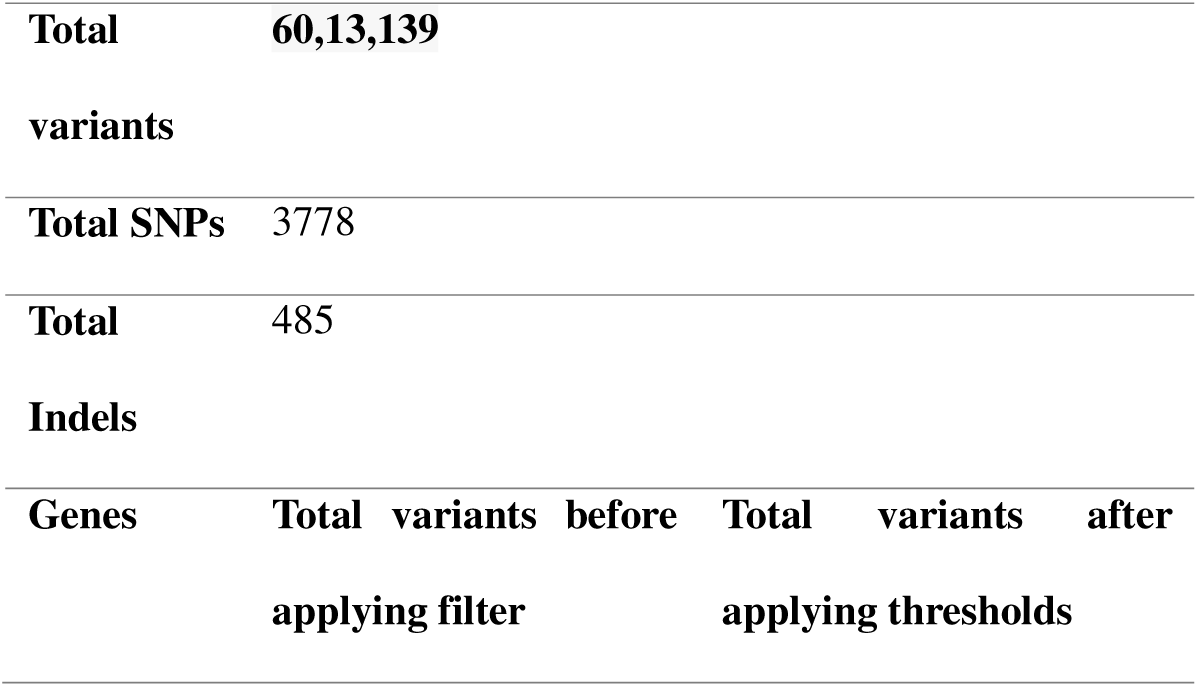
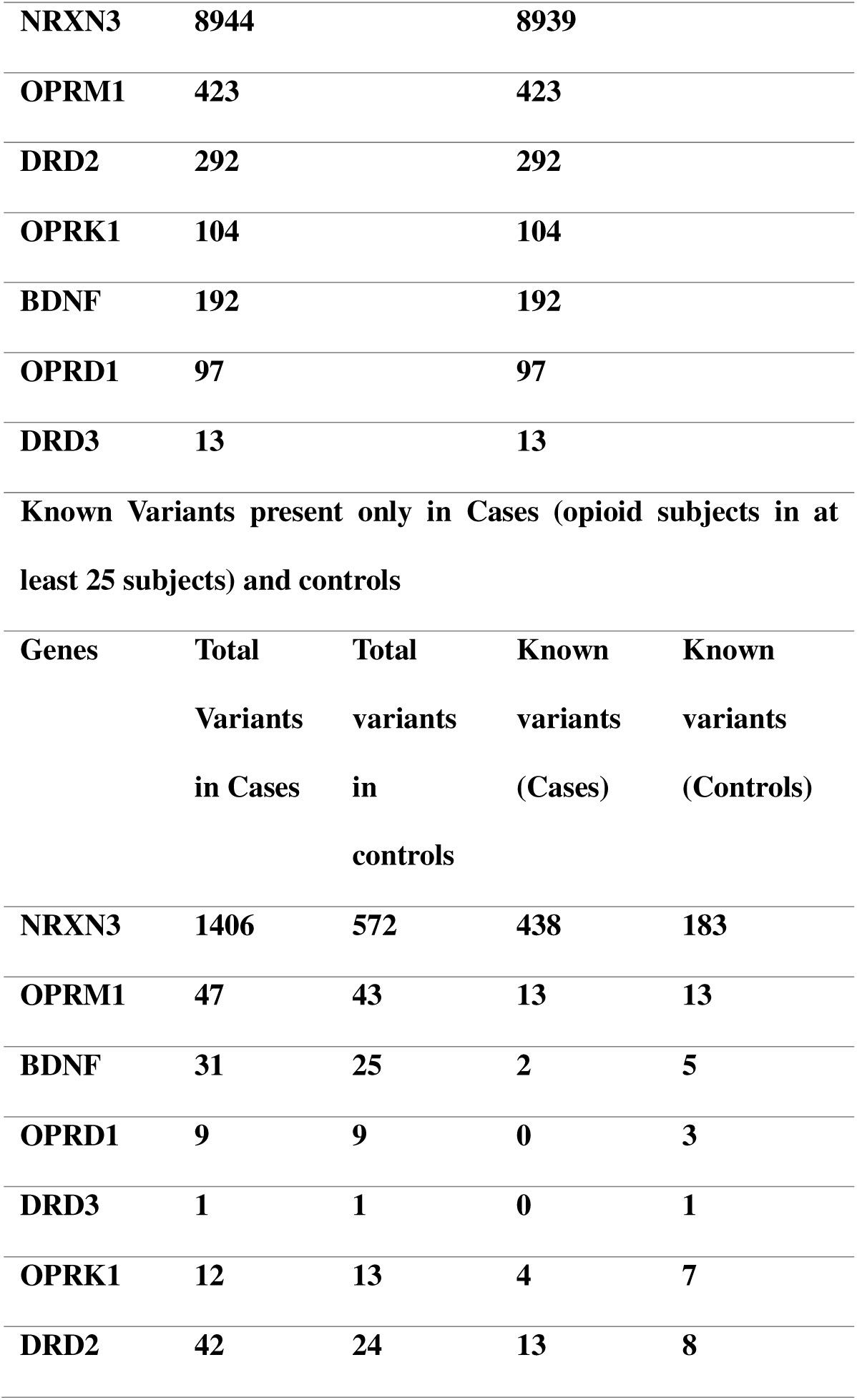
Variants for genes before and after applying quality threshold filters.

**Fig. I.**
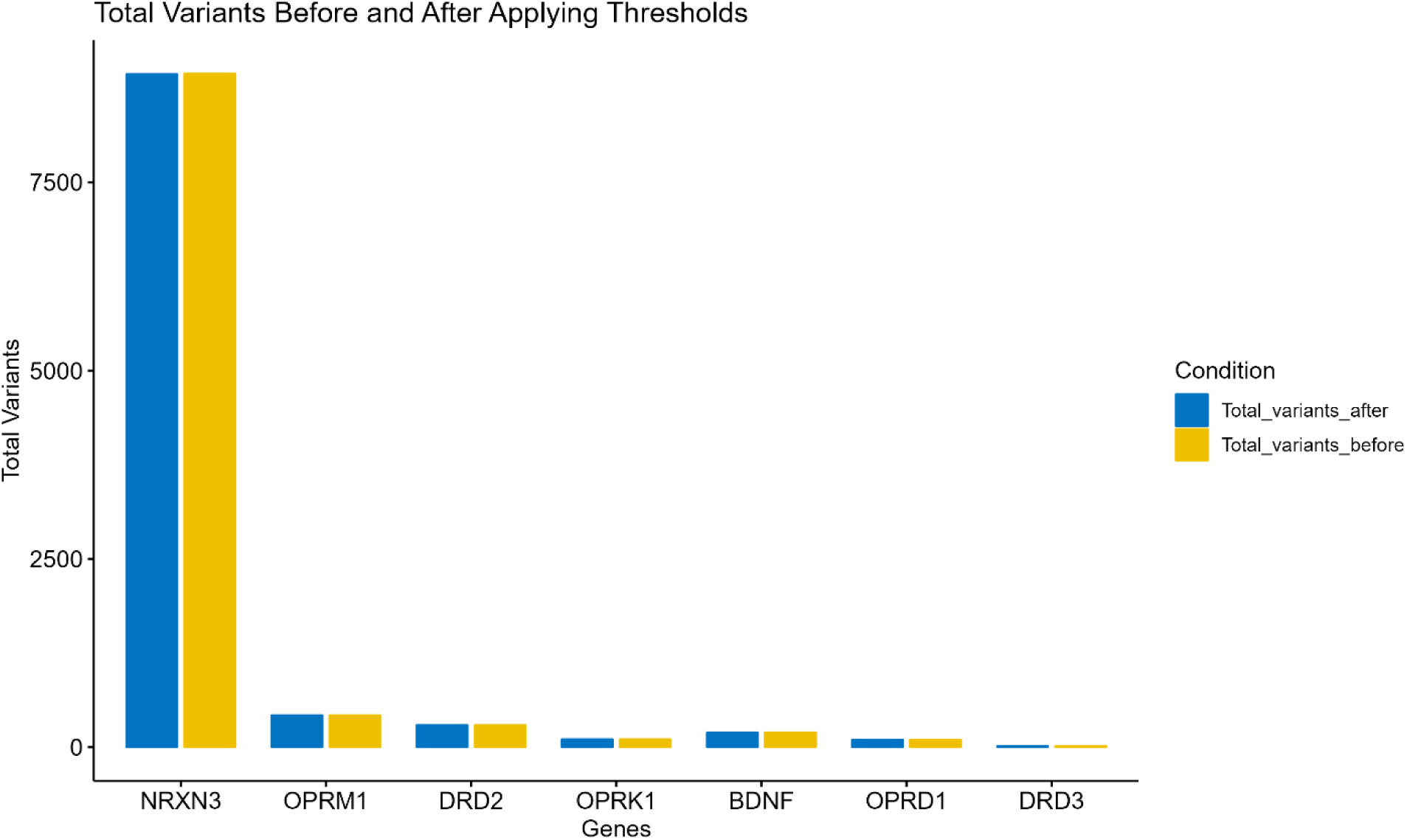
**The total number of variants before and after applying thresholds reveals that the gene NRXN3 exhibits the highest number of variants in both conditions. Conversely, the gene DRD3 shows the lowest number of variants**.

**Fig. II.**
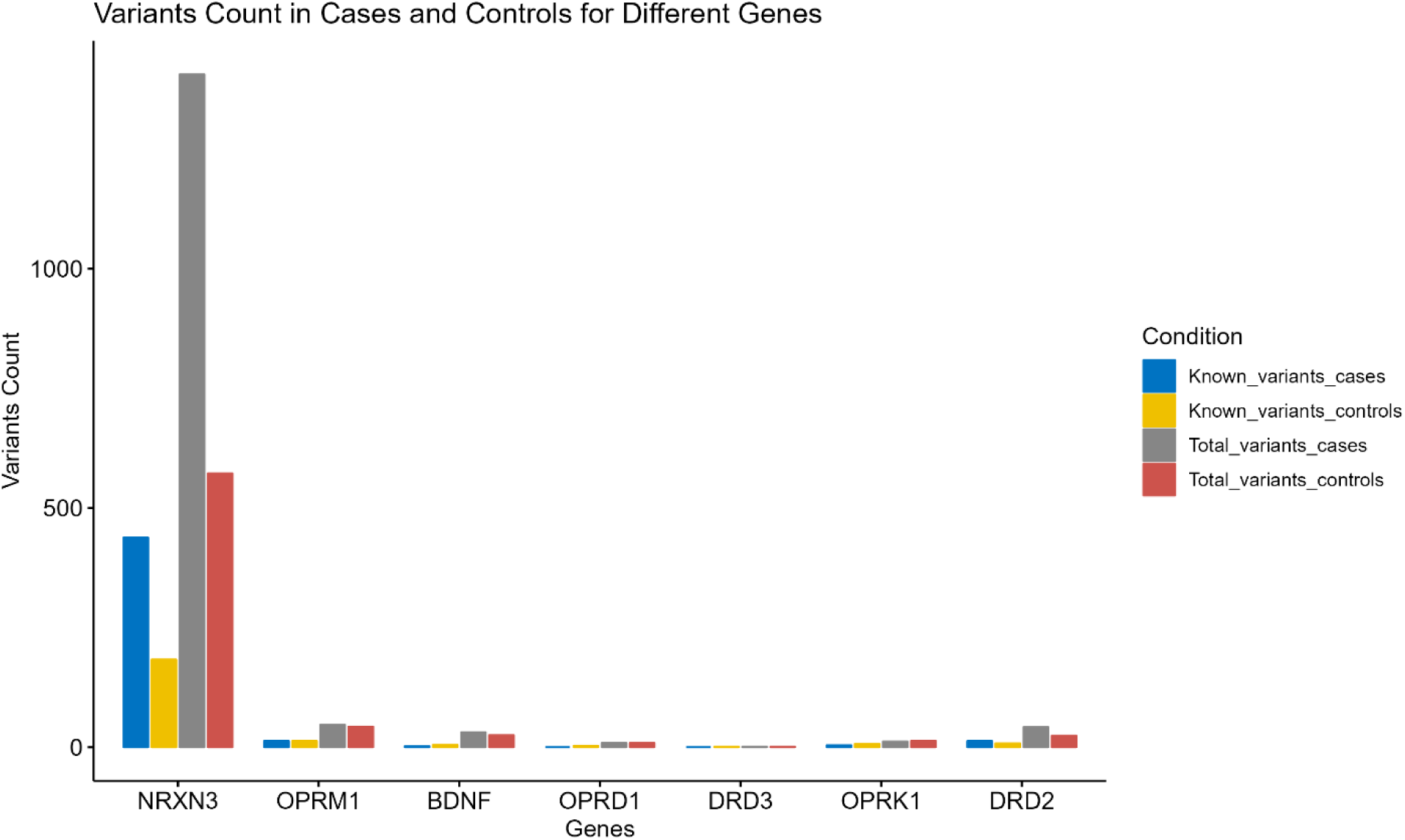
**The bar plot displays the count of known variants in both cases and controls, as well as the total variants in both groups. The NRXN3 gene consistently exhibits the highest number of variants**.

### OPRM1 variants profile

Within the OPRM1 gene located on chromosome 6, our investigation unveiled a total of 47 variations with 13 known mutations, including two Insertion/Deletion variants (INDELs) and various Single Nucleotide Variations (SNVs). The spectrum of these variations encompassed diverse variant types, comprising 3 prime UTR variants, an intron variant, a downstream gene variant, an upstream gene variant, and a non-synonymous variant concurrently exhibiting a stop gained mutation, thereby classified as a HIGH IMPACT mutation. Notably, all mutations, except for one instance of the rs34427887 variant, manifested a heterozygous nature. In contrast, rs34427887 displayed both homozygous and heterozygous manifestations of the stop-gained mutation. Utilizing Gene Set Enrichment Analysis (GSEA) with annotations sourced from the MSigDb database, our findings consistently associated all identified variations with the Hallmark Reactive Oxygen Species Pathway. Concerning the rs34427887 variant, identified with a stop gained mutation, the Clinical Disease Database in ClinVar underscored its correlation with Tramadol response. This clinical insight implies the variant’s impact on drug response rather than a specific disease, emphasizing its role in pharmacological responses. The rs34007356 variant exhibited characteristics of a retained intron. Notably, there were 34 novel variants, including 24 3 prime UTR variants, one missense variant, 5 intron variants, and 3 downstream gene variant and upstream gene variant instances. The Impact of all these variants was classified as MODIFIER, except for one with a MODERATE impact due to a missense mutation resulting in an amino acid change from glutamine to glycine. Intriguingly, all these novel variants exhibited the same GSEA pathway of Hallmark Reactive Oxygen Species Pathway as that observed in known variants. **Fig. III** shows the substitution of SNPs for this gene. The maximum number of substitutions can be seen from A>G and C>T.

**Fig. III.**
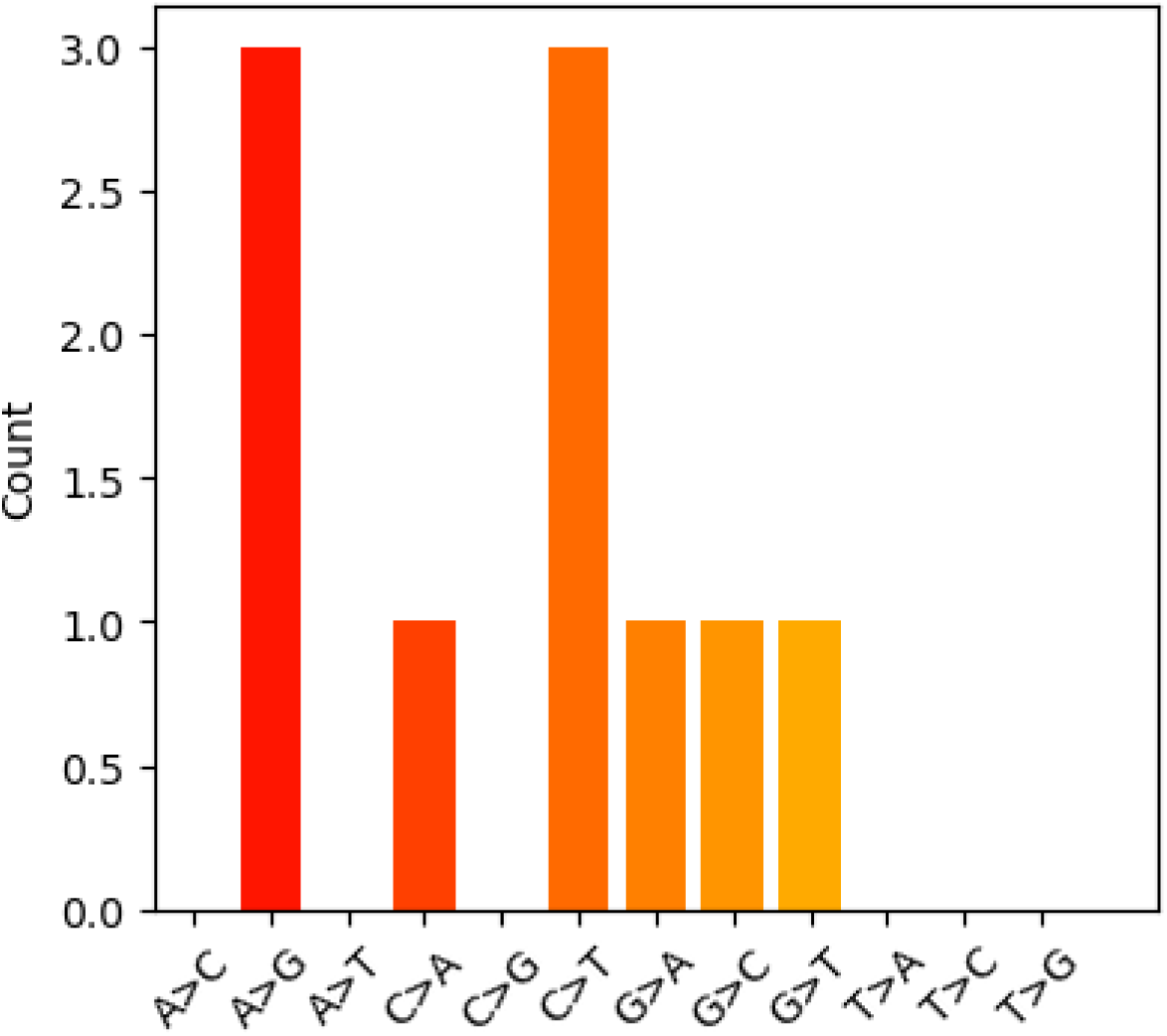
**The figure shows the number of substitutions from the reference allele to the alternate allele for the gene OPRM1**.

### DRD2 variants profile

In the investigation of the DRD2 gene on chromosome 11, 13 variants were identified, consisting of one insertion-deletion (INDEL) and 12 single nucleotide variants (SNVs). All observed variants were manifested as heterozygous genotypes. The majority of variants were categorized with the consequence of "MODIFIER," except for one variant characterized with "MODERATE" impact, denoting a missense variant, and two variants with low impact. Notably, the variant, rs114403746, identified a non-synonymous change at exon 4, substituting guanine with thymine at position 414, leading to an amino acid alteration from methionine (M) to isoleucine (I). This variant was classified as of uncertain significance by Intervar automated analysis and predicted as deleterious by multiple tools including PolyPhen, LRT, MutationTaster, and ClinPred within ClinVar. Additionally, 29 novel variants were discovered, characterized as upstream_gene_variant, downstream_gene_variant, and intron_variant, all attributed to "MODIFIER" impact. Interestingly, one variant of MODIFIER impact had a mutation in the exonic region and, was non-synonymous, replacing guanine(G) with thymine(T) at nucleotide position 414. Among these novel variants, two were annotated as pseudogenes by SnpEff, while two others were identified as belonging to the biotype of retained_intron. The remaining novel variants were identified as protein-coding. Utilizing the bcftools query, a total of 1012 SNPs and 392 INDELs were identified within the examined dataset. **Fig. IV** shows the maximum number of SNPs of G>A and equal numbers of C>T and G>T. **Table. III** shows the variant category in the different regions in the genome.

**Fig. IV.**
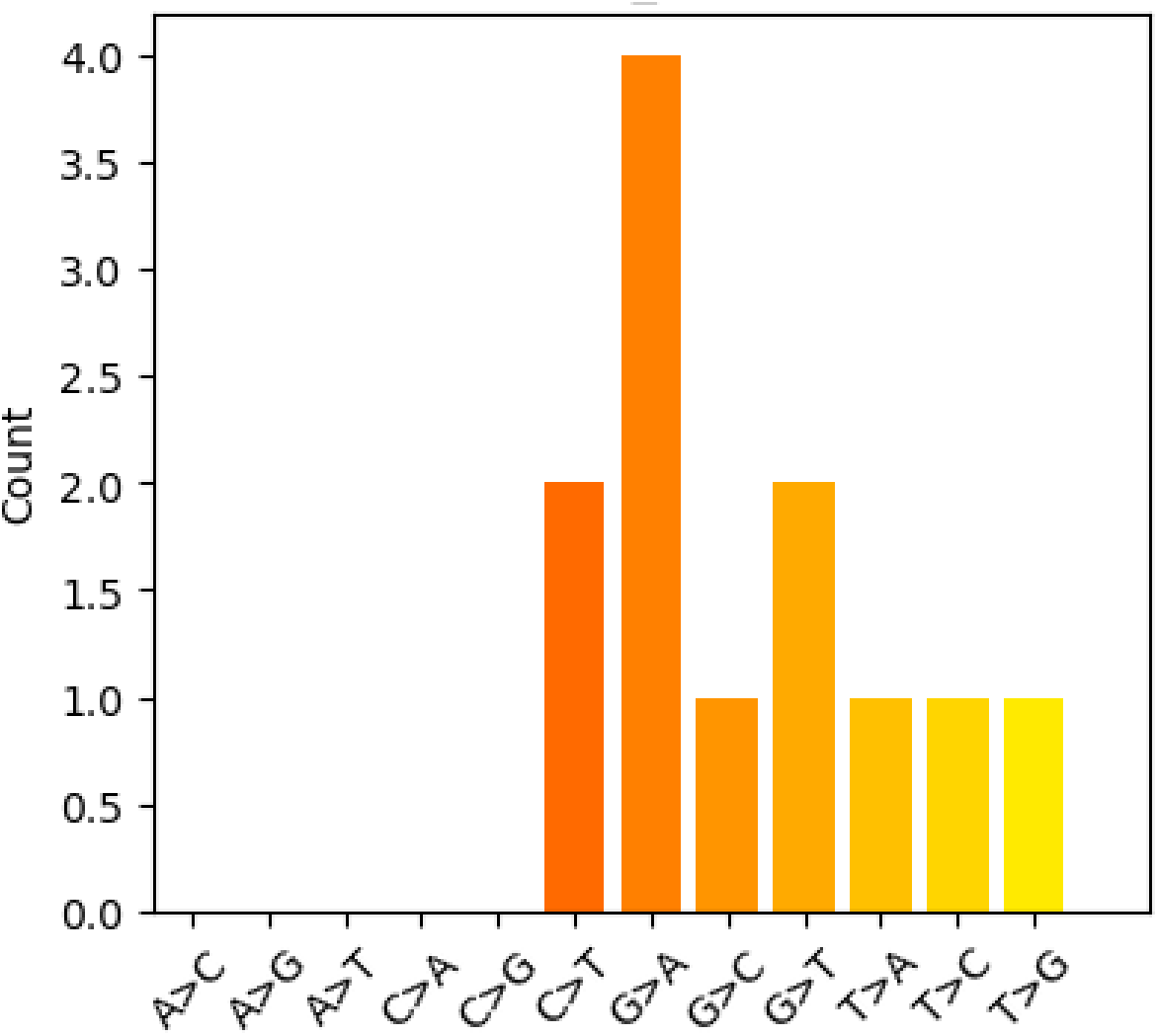
**The figure shows the number of substitutions from the reference allele to the alternate allele for the gene DRD2**.

**Table. II.**
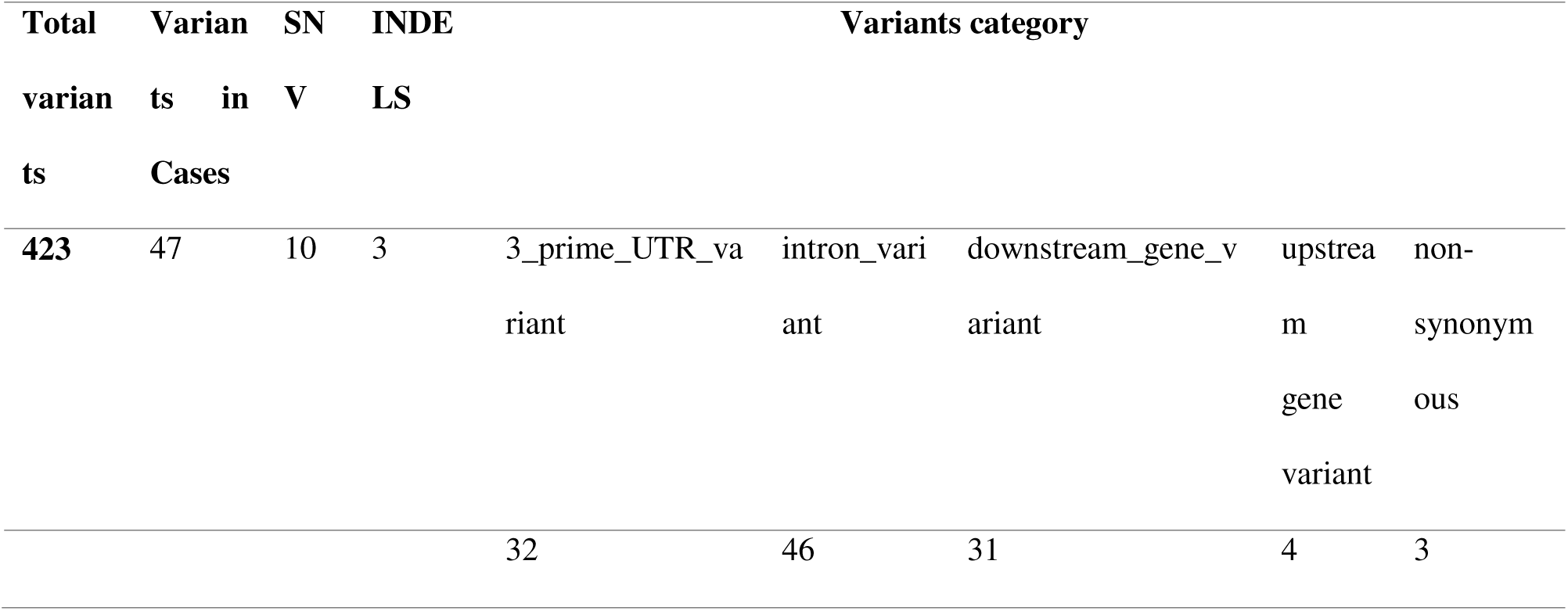
Variants Category based on their genomic region presence.

**Table. III.**
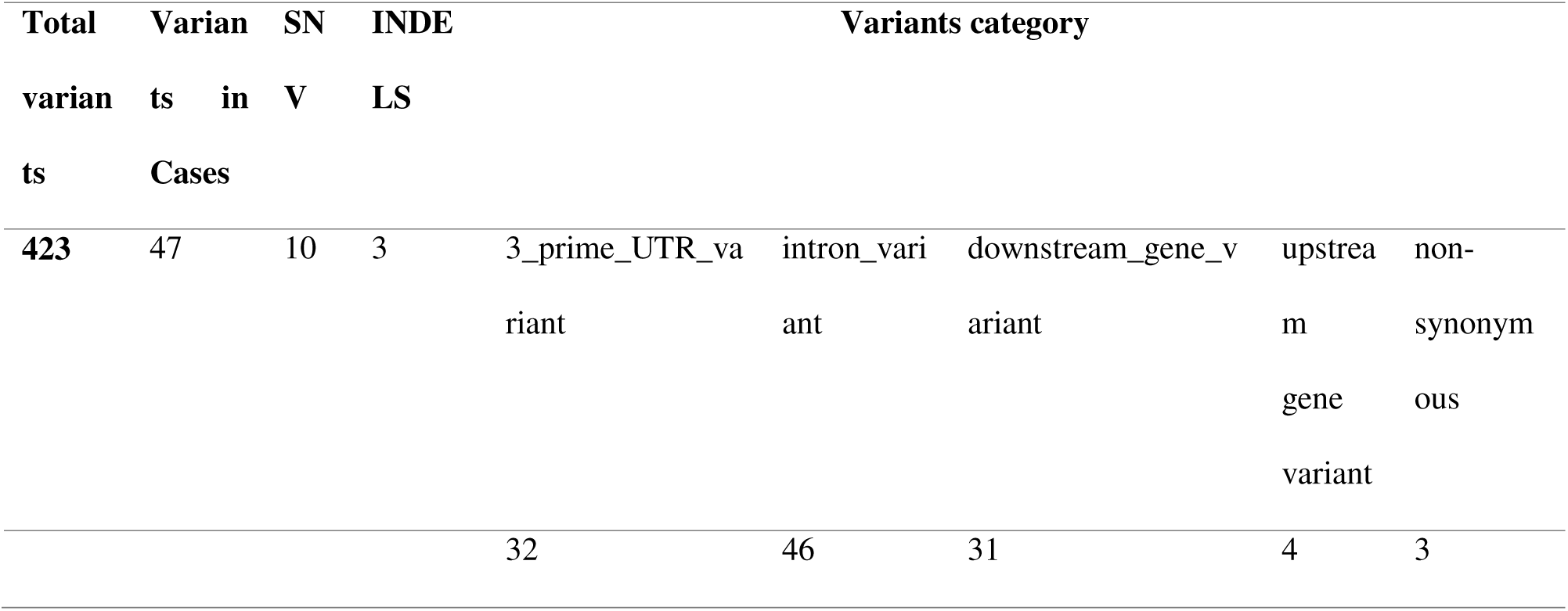
Variants category based on different regions in the genome for DRD2.

### NRXN3 variants profile

Variants within the NRXN3 gene locus on chromosome 14 were extensively analyzed. Out of the total 8939 variants obtained post-threshold filtration, 1406 were observed in case subjects. Among these, 438 variants were annotated with known dbSNP rsIds, while the remaining 968 were classified as novel variants. The variants were categorized into four biotypes: protein_coding, pseudogene, processed_transcript, and retained_intron. Using the bcftools query, it was determined that among the known variants, 400 were single nucleotide variants (SNVs) and 37 were insertions/deletions (INDELs). These variants encompassed various types, including downstream_gene_variant, intron_variant, upstream_gene_variant, missense_variant, 5_prime_UTR_premature_start_codon_gain_variant, and 3_prime_UTR_variant. One notable finding was the presence of a 5_prime_UTR_premature_start_codon_gain_variant at position 72451988 with rsId rs1022111304, albeit with a low impact. Retained introns emerged as a significant category of variants, with 170 instances identified in the known variants. No variants were classified as having a HIGH impact mutation. However, one missense variant was identified with rs2098424152 of MODERATE impact nature changing from serine to isoleucine at position 187. Functional impact predictions revealed that the majority of variants were intron_variant type (419), followed by downstream_gene_variant (28), and 3_prime_UTR_variant (17). The distribution of variants included both homozygous and heterozygous mutations, with the latter being more prevalent. Overall, the analysis highlighted the diverse nature of variants within the NRXN3 gene and their potential implications in molecular functions and disease mechanisms. **Fig. V** shows the maximum number of substitutions for SNPs of A>G. **Table. IV** shows the variation profile for NRXN3.

**Fig. V.**
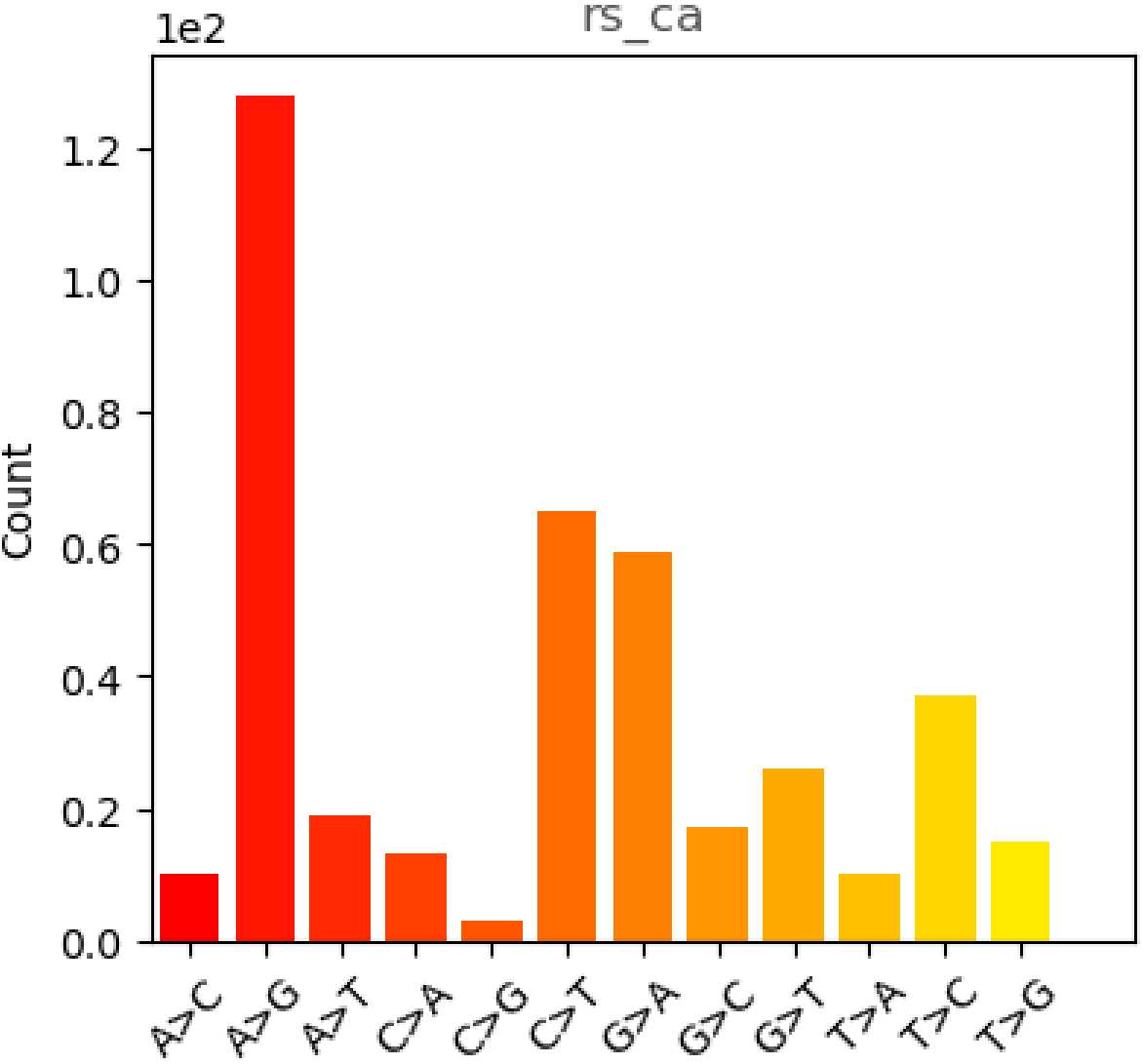
**The figure shows the number of substitutions from the reference allele to the alternate allele for the gene NRXN3**.

**Table. IV.**
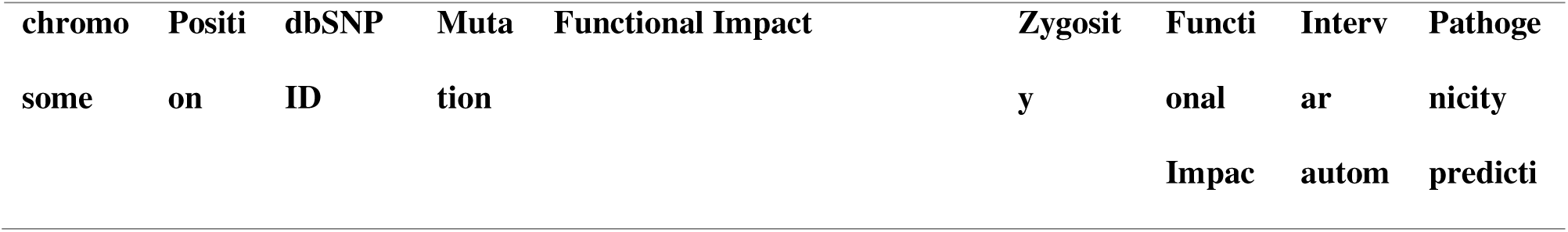
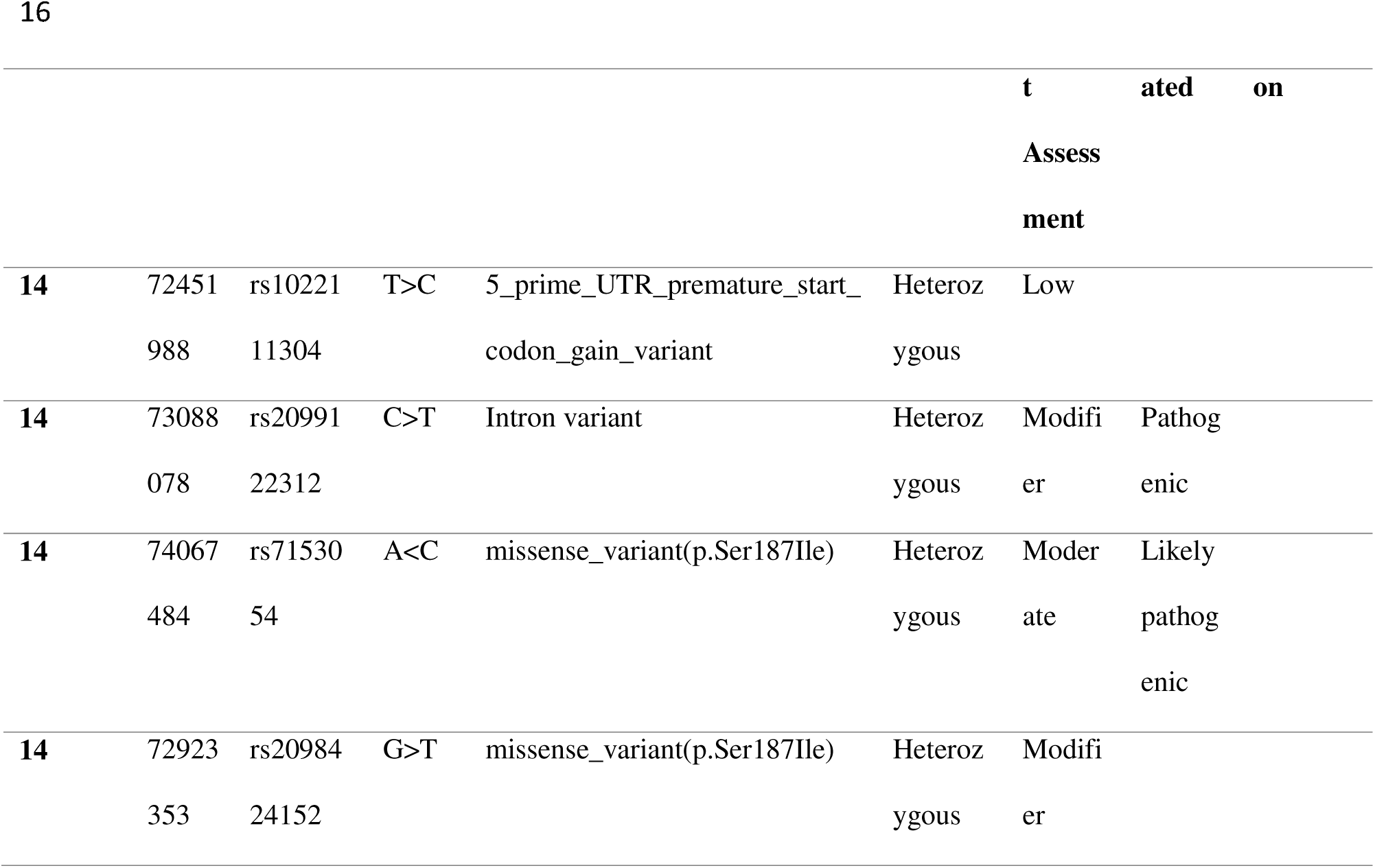
Variants profile for NRXN3 gene.

### BDNF

Using a defined threshold as mentioned, 192 variants were identified within this gene on chromosome 11. Out of the 192 variants, 2 known variants that were SNVs were obtained on chromosome 11. The rest of the novel variants were 3_prime_UTR and upstream_gene_variants. And the impact of these variants was of MODIFIER type. Out of the 29 novel variants, 3 variants, had HIGH impact mutation, and all three variants had frameshift mutation having loss of function mutation. The frameshift had a change in amino acid from in Arginine at position 307, an amino acid change in serine at position 227, and tyrosine at position 172. The gene ID for these frameshift variants was CHM13_G0008566, with 17 transcripts affected having a total transcript affected percentage of 0.88. For novel variants as well as the known ones, the MsigDB showed the Hallmark epithelial-mesenchymal transition and Hallmark UV response pathways. All variants both in known and novel variants had heterozygous mutation. The nature of variants detected in novel variants were 12 3_prime_UTR_variants, 3 frameshift variants and 3 intron variants, 3 splice_donor_variants & Intron_variant, 5 downstream_gene_variant, and 1 splice_region_variant & non_coding_transcript_exon_variant. The 3 prime UTR variants, intron variants, and downstream gene variants showed MODIFIER impact. While splice region variants & intron variants showed a LOW impact. **Fig. VI** shows the maximum number of SNPs of C>T and G>C.

**Fig. VI.**
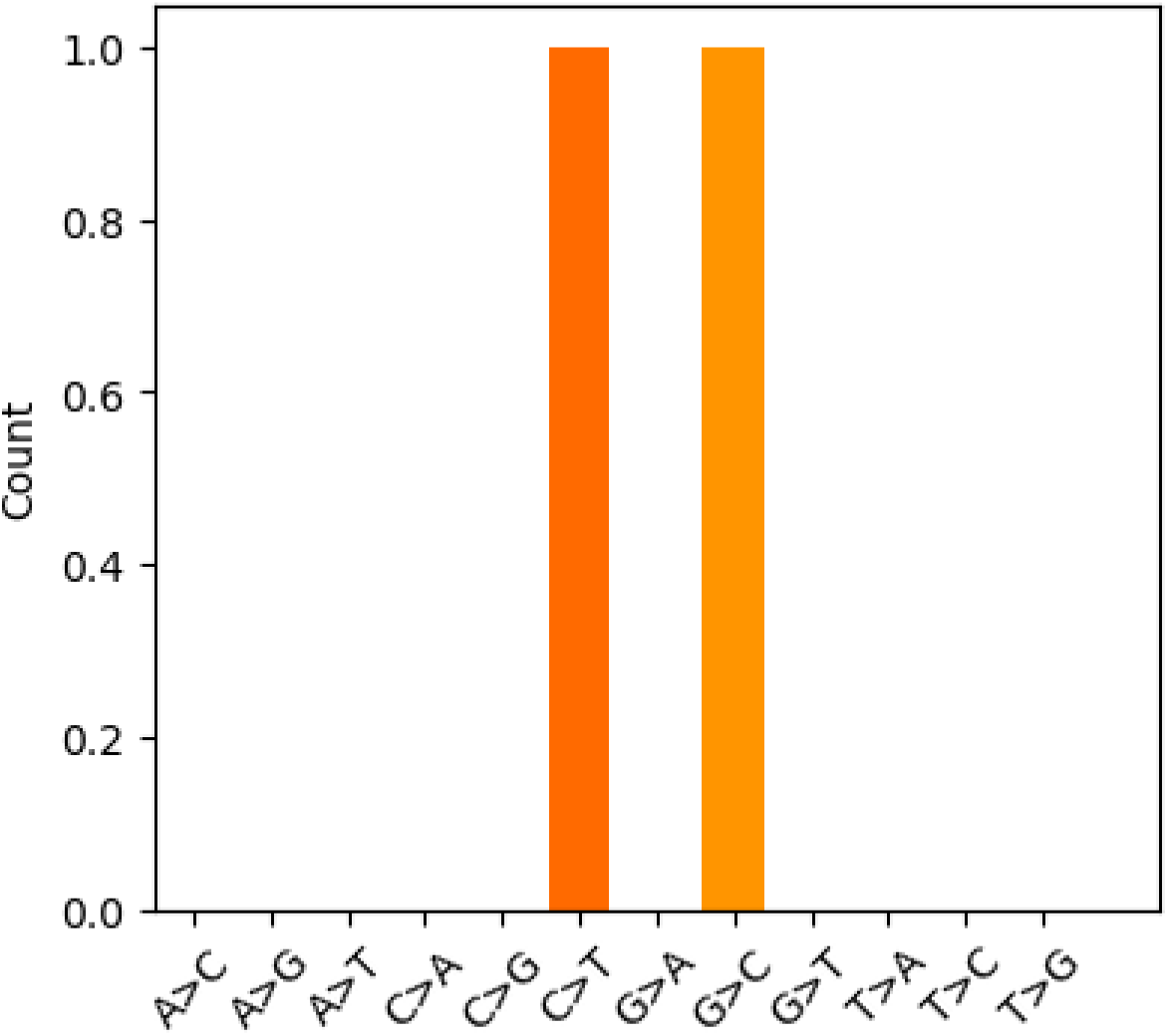
**The figure shows the number of substitutions from the reference allele to the alternate allele for the gene BDNF**.

**Fig. VII.**
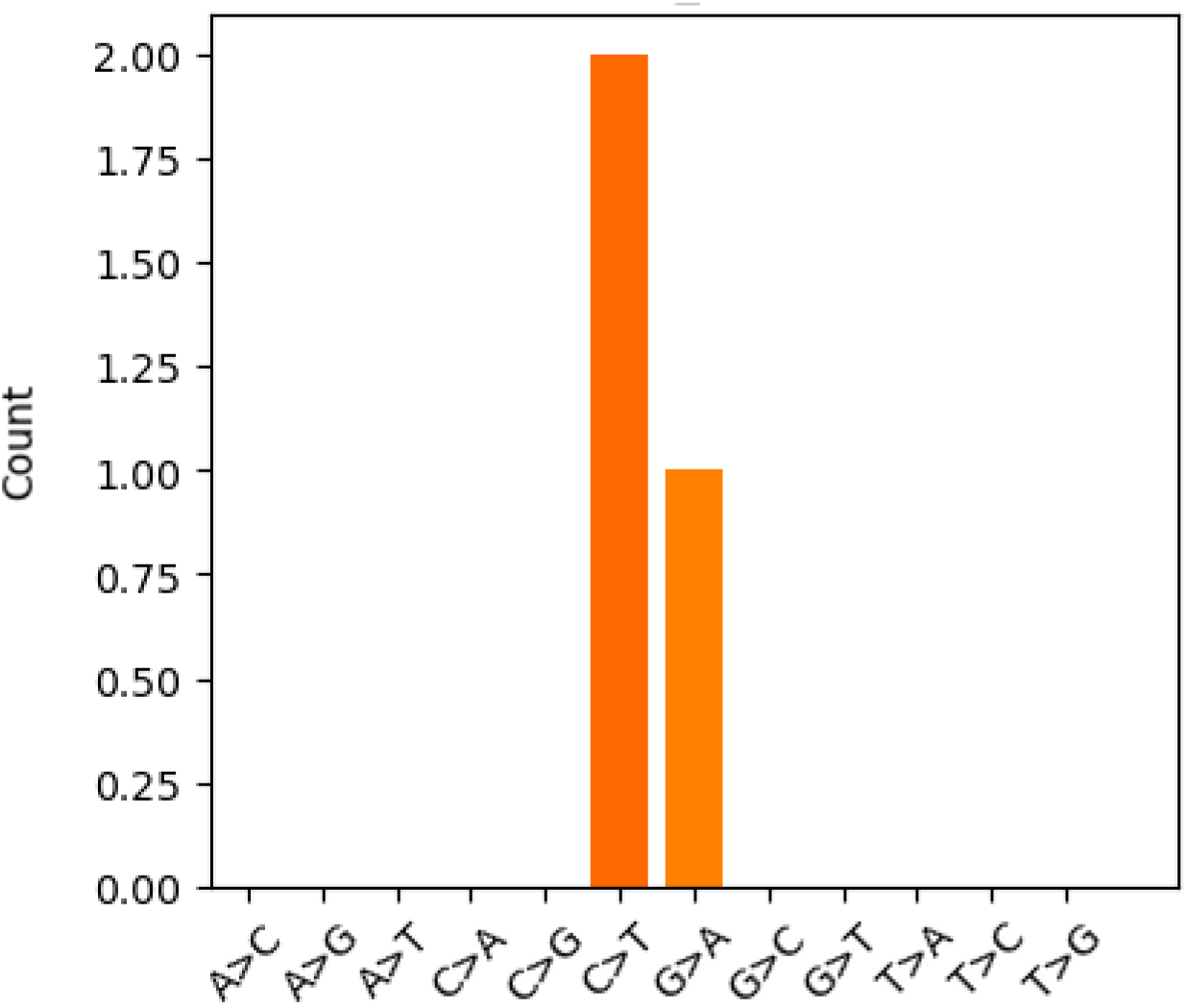
**The figure shows the number of substitutions from the reference allele to the alternate allele for the gene OPRK1**.

### OPRD1 variants profile

All novel variants were observed was this gene that were nine in number with all 3_prime_UTR_variant but one category of downstream gene variant situated on chromosome 1 All these novel variants exhibited a uniform "MODIFIER" impact characterized by heterozygous genotypes.

### DRD3 variants profile

Only one variant was found in case subjects that were novel found on chromosome 3. The type of variant was found to be 3_prime_UTR_variant. The impact of this gene was MODIFIER. Thi mutation was heterozygous.

### OPRK1 variants profile

For OPRK1, the variants were identified on chromosome 8, totaling 12 occurrences exclusively observed in case samples. Among these, 4 variants were recognized as known, while 8 were novel. Within the novel variants, all but one were classified as having MODIFIER impact, except for a single variant characterized by a HIGH impact due to a frameshift mutation affecting leucine at position 348. This frameshift mutation resulted in a loss of function and pertained to the GENE ID CHM13_G0053747, affecting four transcripts, constituting a total proportion of 0.75. Among the known variants, apart from the major MODIFIER, one LOW impact mutation, being synonymous was also identified. For this synonymous variant, the alteration involved a change from valine at position 282. Furthermore, both known and novel variants were annotated to the Hallmark Inflammatory Response pathway. All known variants exhibited heterozygous mutations.

## Discussion

In the study done by [36], high-throughput RNA-sequencing on post-mortem midbrain was done to examine the genome-wide changes in the mid-brain gene expression associated with human opioid abuse from chronic opioid users (N = 30) and drug-free control subjects (N = 20), a described in the Methods [36]. In the current study, we report exploratory variants identified from this gene expression data. Although the original study examined the differentially expressed genes in the mid-brain associated with opioid abuse[36]. We exploited this gene expression data (PRJNA492904) to study the variant type associated with opioid abuse. To our knowledge, this study represents the first attempt to identify variants using Case-Control association analysis from RNA-seq gene expression data of human opioid abusers. We performed variant calling using GATK haplotyecaller and used Snpsift to perform Case-Control association analysis from 30 Cases and 20 Control Cases. We focused on the variants (both homozygous and heterozygous) present exclusively in Cases with chronic opioid use. Variants present in Control samples were excluded from this primary analysis. However, we analyzed the variants in the Control samples to facilitate a comparison between the variants in the Control and Case samples. Our findings indicated that out of the eight (DRD2, DRD3, OPRM1, OPRD1, OPRK1, BDNF, NRXN3, NGFB) genes that were previously associated with human opioid abuse as reviewed by [35], all genes exhibited variants except for NGFB.

The analysis revealed a predominance of heterozygous variants across all genes, mainly located in non-coding regions, suggesting a regulatory role in opioid use disorder (OUD). Additionally, non-synonymous variants in exonic regions were observed for some novel variants, showing deleterious mutations. Notably, a frameshift mutation was identified in the OPRK1 gene, and three frameshift mutations with high impact, as annotated by SnpEff, were found in the BDNF gene. A heterozygous mutation with rs1022111304 in the NRXN3 gene was exclusively present in case samples and absent in controls, highlighting its potential significance. Similarly, the missense mutation in NRXN3 with dbSNP ID rs2098424152 was oberved. These mutations require further analysis in different populations and experimental validation. The maximum number of variants were found in NRXN3. The number of substitutions for genes shows the maximum number of susbstitutions for NRXN3 gene with A>G substitution highest in number. The *NRXN3* gene is one of the largest genes in the human genome containing 1,826,818 base pairs[37]. Neurexin-3-alpha is a protein that in humans is encoded by the *NRXN3* gene[38]. Neurexins (NRXNs) are presynaptic transmembrane proteins that play a role in the development and function of synapses[39], and this gene has also been found to be associated with impulsiveness in patients with substance abuse[40]. We further did annotation of variants associated with genes exhibiting variants to determine the functional impact of variants on gene function. Existing suggestive evidence indicates that polymorphisms associated with NRXN3 induce impulsivity that leads to risky behaviors, that may give rise to behavioral disorders such as addiction[40]. This finding may prove to be significant in understanding the genetic architecture of addiction because impulsivity is an important risk factor for addictions and other behavioral disorders[39]. Evidence is accumulating that NRXN3 may be involved in a pathway that influences vulnerability to addiction in general[39]. In that sense, our findings add to the growing evidence that implicates polymorphism in NRXN3 in risk for substance use problems and/or dependence[41,42]. Given the maximum number of variants found in the NRXN3 gene compared to other genes, our findings suggest that NRXN3 polymorphisms may be modestly associated with specific types of impulsivity and substance use problems. Impulsivity increases the risk for several behavioral disorders such as addictions and therefore is an outstanding candidate trait for further study. A more complete description of the genetic architecture of impulsivity is likely to have a substantial impact on our understanding of the genetic architecture of a variety of behavioral disorders. Furthermore, these patterns may vary among different populations. Findings such as these, despite their limitations, can be critical for advancing our understanding of the molecular mechanisms linking variants associated with genes to behavior. NRXN3 is a promising candidate gene for addiction vulnerability and merits further study. The majority of clinical genetic testing predominantly targets regions of the genome that encode proteins. However, the significant role of variants in non-coding regions in penetrant diseases is increasingly being recognized. Previously, several of the non-coding variants associated with the substance use disorder were identified[43]. The application of whole genome sequencing in clinical diagnostic settings is expanding across a broad spectrum of genetic disorders. Despite this progress, there is currently no guidance on how existing guidelines, primarily designed for variants in protein-coding regions, should be adapted for variants identified in the gene expression region. Due to this reason, we applied variant filtering thresholds to assess differences in variant counts before and after their application. As shown in Table. I, the only notable difference in variant numbers is observed in the NRXN3 gene, with no differences detected in the other genes. Table. I illustrate the differences in variants between Cases and Controls. The only significant difference in variants is observed in the NRXN3 gene. This difference is visualized in Fig. 2, where both total and known variants are prominently displayed for the NRXN3 gene alone. In the NRXN3 gene and other genes analyzed in this study, most variants were located in non-coding regions, specifically of an intronic nature. This finding suggests a potential involvement of intronic variants in opioid use addiction. Several studies now indicate a possible association between intronic variants and various types of addictions Several studies have identified intronic variants implicated in addiction. For instance, [16] found intronic variants associated with heroin addiction. [44] reported a significant association between an intronic variant and treatment outcomes in opioid dependence. Additionally, [45] reported the associations of intronic single nucleotide polymorphisms (SNPs) as a risk factor for internet addiction. These observations aim to increase the number and range of non-coding region variants that can be clinically interpreted, which, together with a compatible phenotype, can lead to new diagnoses and catalyze the discovery of novel disease mechanisms. There is substantial evidence that genetic factors influence an individual’s susceptibility to addiction. However, identifying the specific genes that contribute to this risk remains a challenging and ongoing endeavor. For a given substance use disorder, there is an expectation that there are genes that affect addiction vulnerability to that particular substance, and that there are also genes that influence traits that increase vulnerability to addiction, in general. Our study offers suggestive evidence that the variants in the NRXN3 might act as impulsivity factors as a key construct leading to risky behaviors and ultimately opioid addiction. Further investigation with detailed phenotypic assessment that includes patterns of substance use, substance use disorder diagnoses, multiple measures of impulsivity, and additional NRXN3 polymorphisms should be conducted. We recognize that our study has certain limitations due to the lack of standard guidelines for variant identification from RNA-Seq data and needs to be interpreted with some caution. More detailed assessments of the potential associations between important addiction-related phenotypes and genetic variation across NRXN3 need to be conducted. From the current analysis, we suggest that transcriptomic data can be used for variant identification which is cost-effective and offers the gene expression and variants analysis at the same time.

## Conclusion

Our comprehensive bioinformatics analysis from a case-control study identified some characteristic known and unknown variations in opioid-addicted subjects. Although our study shows characteristic variations in certain genes, an assay comprising multiple biomarkers that are differentially expressed could be attempted in the future. If this is successful, the number of biomarkers developed will depend on their validation in a large cohort of patients and the translation of these findings to clinical practice. Our current study, we believe, is decisive in understanding the significance of variations in opioid addiction as well as the utilization of transcriptomic data for variant analysis. Based on the current SNP analysis results, further study seems warranted to determine the potential association of NRXN3 variants as biomarkers and/or therapeutic targets for substance abuse.

## Data Availability

All data produced are available online at https://www.ncbi.nlm.nih.gov/bioproject/PRJNA492904

## Author Contributions

S.A. designed the analysis, applied methods, and wrote the main manuscript; B.C. reviewed and edited the manuscript; S.K. supervised, edited, and reviewed the manuscript. All authors read and approved the final manuscript.

## Availability of data and materials

A publicly available dataset was analyzed in this study. This data can be found in the link NCBI Sequence Read Archive (SRA; www.ncbi.nlm.nih.gov/sra) under accession number PRJNA492904

## Ethical approval

This declaration is not applicable

## Acknowledgments

The authors acknowledge the support extended by the All India Institute of Medical Sciences (AIIMS), New Delhi for providing computational facilities and Indian Council of Medical Research(ICMR) for providing funding.

